# Peripheral and Cerebral Vascular Reactivity in Black and White Women: Examining the Impact of Psychosocial Stress Exposure Versus Internalization and Coping

**DOI:** 10.1101/2023.03.16.23287388

**Authors:** Zachary T. Martin, Iman O. Al-daas, Natalia Cardenas, John O. Kolade, Emily R. Merlau, Joshua K. Vu, Kyrah K. Brown, R. Matthew Brothers

**Author notes:** **Corresponding author:** R. Matthew Brothers, R. Matthew Brothers, PhD, Department of Kinesiology, College of Nursing and Health Innovation, University of Texas at Arlington, 500 W. Nedderman, MAC 147, Arlington, TX 76019 USA.

## Abstract

Black women have the highest rates of cardiovascular and cerebrovascular disease prevalence and mortality in part due to blunted vascular function. Psychosocial stress likely also contributes but its relationship to vascular function remains incompletely understood. Recent studies suggest that internalization and coping strategies are more important than stress exposure alone. We hypothesized that Black women have blunted peripheral and cerebral vascular function and that, among Black women, this would be inversely related with psychosocial stress internalization/coping but not stress exposures. Healthy Black (*n* = 21; 20 ± 2 yr) and White (*n* = 16; 25 ± 7 yr) women underwent testing for forearm reactive hyperemia (RH), brachial artery flow-mediated dilation (FMD), and cerebrovascular reactivity (CVR). Psychosocial stress exposure (adverse childhood experiences, ACEs; past week discrimination, PWD) and internalization/coping techniques (John Henryism Active Coping Scale, JHAC12; Giscombe Superwoman Schema Questionnaire, G-SWS-Q) were assessed. RH and CVR (*p* > 0.05) were not different between groups whereas FMD was lower in Black women (*p* = 0.007). Neither ACEs nor PWD were associated with FMD in either group (*p* > 0.05 for all). JHAC12 scores were negatively associated with FMD in Black women (*p* = 0.014) but positively associated with FMD in White women (*p* = 0.042). SWS-Succeed was negatively associated (*p* = 0.044) and SWS-Vulnerable tended to be negatively associated (*p* = 0.057) with FMD in Black women. These findings indicate that blunted FMD in Black women may be due more to internalization and maladaptive coping than stress exposure alone.

## Introduction

Among women in the United States, Black women suffer from the highest rates of hypertension, coronary artery disease, stroke, and total cardiovascular disease (CVD) mortality (1). Importantly, peripheral and cerebral vascular dysfunction precede overt CVD and are predictive of CVD risk and mortality (2-6). We and others have reported blunted peripheral and cerebral vascular function in Black men and women (7-14). However, this is not always a consistent finding (7, 15) and, alarmingly, Black women are underrepresented in this literature despite have similarly high rates of disease risk/prevalence as Black men (1). Furthermore, although these investigations have been critical for identifying race/ethnicity-related disparities in vascular function, more work is needed to better understand the influence of upstream factors, such as social determinants of health (SDOH), on cardiovascular health and subsequent disease risk in Black women (16).

SDOH range from access to quality healthcare and education to employment opportunities and living in a safe environment (16). In the United States, structural racism (e.g., policies that systematically disenfranchise racial/ethnic minoritized groups) has put many Black Americans at a social disadvantage (16), which ultimately has numerous negative impacts on all aspects of life, including cardiovascular health (16). Indeed, Black Americans are more likely to experience adverse childhood experiences (17) as well as interpersonal racism (18, 19). Accordingly, constant psychological stress from adverse social conditions causes biological responses (i.e., allostasis), which, over time, cause neurohormonal imbalances, including augmented systemic inflammation and oxidative stress (i.e., allostatic overload) (20, 21). These alterations can subsequently impair vascular responsiveness/health (22, 23); however, the precise psychophysiological pathways and mechanisms remain incompletely understood (16). Interestingly, a growing literature suggests that internalization of stressful events and stress coping techniques (e.g., Superwoman Schema or John Henryism (24, 25)) may be more important to cardiovascular health and function than exposure to psychosocial stressors alone (26-29). Therefore, we investigated the relationship between vascular function and psychosocial stress exposure versus internalization and coping. We hypothesized that 1) peripheral and cerebral vascular function would be blunted in Black relative to White women and 2) among Black women, internalization and maladaptive coping would exhibit a stronger negative relationship with vascular function than exposure to stressful experiences alone.

## Materials and Methods

### Ethical Approval

Ethical approval was obtained from the University of Texas at Arlington Institutional Review Board (#2021-0270 and #2022-0541). All participants were informed of the experimental procedures and risks before participating. Verbal and written informed consent was obtained from each participant. The experimental protocols aligned with the guidelines set forth by the Declaration of Helsinki except for registration in a database.

### Participants

Thirty-eight Black (*n* = 21) and White (*n* = 17) women were recruited from the Dallas-Fort Worth Metroplex via flyers, social media, and word-of-mouth from 2021 through 2023. Participants were included if they self-reported being of Black or White race/ethnicity, women, and age 18-40 yr. Participants were excluded if they reported a history of smoking within 2 years or being a current smoker; taking vasoactive medications; or having overt cardiovascular, metabolic, or neurological disease. All data from one White participant were excluded due to persistent COVID-19 symptoms, which have been reported to have negative vascular effects (30), leaving a total of 37 participants (*n* = 21 Black women and *n* = 16 White women).

### Study Design and Protocol

During a single visit to the laboratory, participants completed a routine venous blood draw, social determinants of health questionnaires, and peripheral and cerebral vascular function testing. Testing occurred following a minimum 8 hr fast, at least an 8 hr abstention from caffeine and medications/supplements, and at least a 24 hr abstention from alcohol and vigorous exercise.

### Measurements

#### Initial Measurements

After participants provided informed consent and completed the health history questionnaire, height and weight were measured using a stadiometer and scale (Seca 769; Seca North America; Chino, CA, USA). Body composition was further assessed as the waist-to-hip ratio following standard guidelines (31).

#### Venous Blood Draw

A fasting venous blood sample was taken to assess glucose, lipid, estradiol, and progesterone concentrations along with HbA1c (Labcorp; Burlington, NC, USA). To assess circulating concentrations of C-reactive protein (CRP), a marker of inflammation, blood was collected in a serum separator tube, centrifuged, and analyzed via an immunochemiluminometric assay for CRP (Labcorp; Burlington, NC, USA).

#### Questionnaires

##### Participant Characteristics

To assess each participant’s self-reported physical activity habits, the International Physical Activity Questionnaire (IPAQ) (32) was administered. Due to the relatively young age of both cohorts, we assessed an index of childhood socioeconomic status by asking participants about the highest level of education achieved by their mother and father (33, 34). Seven educational categories were provided, ranging from “Less than high school” to “Master’s degree/terminal professional degree,” and assigned a value from 1-7, respectively (24, 34).

##### Psychosocial Stress Exposure

We used two validated questionnaires to assess exposure to psychosocial stress. To assess early life adversity, which is associated with decrements in mental and physical health across the lifespan (35, 36), we administered the Adverse Childhood Experiences (ACEs) questionnaire (37). ACEs (0-10 experiences) were summed to provide an overall ACEs score. In order to assess an index of short-term psychosocial stress, we administered the Past Week Discrimination (PWD) questionnaire, a ten-item survey inquiring about the frequency (0-3+) of race/ethnicity-related experiences of discrimination during the past 7 days (38). The number of exposures to experiences of discrimination were then summed, yielding an overall PWD score ranging from 0-30, with higher values representing a greater degree of stress exposure.

##### Psychosocial Stress Internalization and Coping

High effort coping with life’s obstacles in the face of limited resources was measured via the John Henryism Active Coping Scale (JHAC12), a 12-item questionnaire that asks the participant to state the extent (from completely false to completely true) to which statements such as “I don’t let my personal feelings get in the way of doing a job” are true for them (39). Responses were coded 1-5, respectively, and summed, thereby providing an overall JHAC12 score (39). Woods-Giscombe, et al. developed the Giscombe Superwoman Schema Questionnaire (G-SWS-Q), which operationalizes the Superwoman Schema (SWS) Conceptual Framework to quantify psychological stress processes specifically associated with being a Black woman (24, 40). Therefore, to measure stress coping strategies and SWS internalization, Black participants completed the G-SWS-Q, which is a 35-item instrument designed to measure the cognitive, affective, and behavioral underpinnings associated with being a Strong Black Woman in the United States (24, 40). Responses were scored according to the developer’s instructions, and provided an overall SWS score as well as scores for 5 subscales: 1) Obligation to present an image of strength; 2) Obligation to suppress emotions; 3) Resistance to being vulnerable; 4) Intense motivation to succeed; and 5) Obligation to help others (24). Higher scores indicated greater endorsement of the various SWS constructs.

#### Brachial Artery Flow-Mediated Dilation & Forearm Reactive Hyperemia

Following 15 min of supine rest, measures of microvascular and macrovascular function were performed as previously described (41) and in accordance with the latest evidence-based recommendations (42). Briefly, the right arm was abducted and supported at heart level while a pneumatic cuff was wrapped around the forearm ∼1 cm distal to the antecubital fossa. The brachial artery was insonated ∼5-10 cm proximal to the antecubital fossa via an adjustable frequency (10-13 MHz) linear array Doppler ultrasound probe (LOGIQ P5; GE Healthcare; Chicago, IL, USA), held in place by a stereotactic probe holder at an angle of 60°. B-mode images of the brachial artery were optimized to ensure delineation between the vessel wall and lumen during offline analysis. The probe was then set to duplex mode (pulsed frequency of 5 MHz) for simultaneous imaging of the artery and blood velocity, with the sample volume set to include the entire artery without extending beyond its wall. Brachial artery diameter (*D;* cm) and blood velocity (*V*_blood_; cm/s) were continuously recorded throughout a 2 min baseline period, after which the cuff was inflated to 220 mmHg to occlude circulation to the forearm for 5 min (Rapid Cuff Inflation System; D. E. Hokanson, Inc.; Bellevue, WA, USA). Upon cuff release, *D* and *V*_blood_ were recorded for a 3 min recovery period. Digital video of the entire protocol was recorded and saved (Elgato/CORSAIR; Fremont, CA, USA) for later analysis using a commercially available edge-detection and velocity tracking software (FMD Studio; Quipu; Pisa, Italy). Brachial artery flow-mediated dilation (%FMD) was defined as ((*D*peak − *D*baseline)/ *D*baseline) × 100. Angle-corrected *V*_blood_ was calculated as the full area of the Doppler envelope each second and averaged between anterograde and retrograde velocity (*V*_mean_). Reactive hyperemia (RH) was calculated as the peak as well as the relative (absolute Δ and percentage) increase in blood velocity following cuff release. Shear rate, as s^-1^, was calculated as 4 × *V*mean/*D* and shear rate AUC was calculated as the sum of the second-by-second shear rate above baseline up to *D*_peak_.

#### Cerebral Vascular Function

Cerebral vascular function was assessed as the cerebral vasodilatory reactivity to steady-state hypercapnia as previously described in detail by our lab and others (6, 43-45). While supine, heart rate was monitored via ECG (CardioCard; Nasiff Associates; Central Square, NY, USA) and brachial artery blood pressure was measured via an oscillometric electrosphygmomanometer (Tango M2; SunTech Medical; Morrisville, NC, USA). The partial pressure of end-tidal CO_2_ (P_ET_CO_2_) and peripheral oxygen saturation (SpO_2_) (Capnocheck® Plus; BCI/Smiths Medical; Minneapolis, MN, USA) along with respiratory rate (Pneumotrace II; UFI; Morro Bay, CA, USA) were also continuously measured. Continuous beat-to-beat blood pressure was monitored via finger photoplethysmography (Finometer PRO; Finapres Medical Systems; Enschede, Netherlands). Left middle cerebral artery blood velocity (MCAv) was measured using a 2 MHz probe (TOC Neurovision; Multigon Industries; Yonkers, NY, USA) fixed at the temporal window with a headband. The participants were then fitted with a mouthpiece and 3-way stopcock valve (Hans Rudolph, Inc.; Shawnee, KS, USA) that led to either ambient air or a 5 L rubber bag (GPC Medical Ltd.; New Delhi, Delhi, India) pre-filled with a 6% CO_2_ gas mixture (21% O_2_, nitrogen balance). Baseline data were collected for 3 min while the participant breathed ambient air, after which the valve was switched so that the participant began breathing air from the bag, which was continuously supplied with 6% CO_2_. Following 3 min of steady-state hypercapnia, the valve was switched back to ambient air and data collection continued for a 3 min recovery period. Data from the last minute of baseline and the last minute of hypercapnia were selected and used in the analysis. Cerebrovascular conductance index (CVCi) was calculated as *CVCi* = *MCAv*/*MAP*, and cerebrovascular reactivity was assessed as *ΔMCAv*/*Δ PETCO*2, *ΔCVCi*/*Δ PETCO*2, and %*CVCi*/*Δ PETCO*2.

### Statistical Analysis

All data were processed using GraphPad Prism 9 (v. 9.4.0; GraphPad Software, LLC; San Diego, CA, USA) and SAS 9 (v. 9.4; SAS Institute; Cary, North Carolina, USA). The data were assessed for normality via the D’Agostino-Pearson normality test. Group comparisons of participant characteristics and outcomes were evaluated via Welch’s *t*-test, the Mann-Whitney *U* test, and ANCOVA. Preliminary analyses revealed that 1) brachial artery FMD was the only vascular outcome indicating differences between groups and 2) there were no discernible relationships between psychosocial stress outcomes and vascular outcomes when collapsing the groups. Therefore, we performed Spearman correlation analyses between the psychosocial stress variables and brachial artery %FMD in Black and White women separately. RH, FMD, and CVR data from two Black women and one White woman were deemed statistical outliers via Grubbs’ test and excluded from the analysis. All data are presented as mean ± SD or individually along with the group mean, and the level for statistical significance was set *a priori* at α = 0.05.

## Results

### Participant Characteristics

Participant characteristics are provided in Table 1. There were no differences between groups in BMI, waist-to-hip ratio, resting heart rate and blood pressure, physical activity and sedentary levels, nor childhood socioeconomic status as indexed by parental education level (*p* > 0.05 for all). However, Black women were younger than White women (*p* = 0.035).

**Table 1.** Participant Characteristics.

| | Black Women<br>mean ± SD (*n* = 21) | White Women<br>mean ± SD (*n* = 16) | *p* value |
| --- | --- | --- | --- |
| Age (years) | 20 ± 2 | 25 ± 7 | 0.035 |
| BMI (kg/m<sup>2</sup>) | 24.5 ± 4.3 | 24.8 ± 2.7 | 0.796 |
| Waist-to-hip ratio (au) | 0.78 ± 0.05 | 0.74 ± 0.05 | 0.061 |
| Heart rate (bpm) | 62 ± 7 | 64 ± 10 | 0.419 |
| Systolic blood pressure (mmHg) | 114 ± 7 | 117 ± 6 | 0.354 |
| Diastolic blood pressure (mmHg) | 73 ± 5 | 73 ± 6 | 0.883 |
| Physical activity (MET-min/week) | 3795 ± 3428 | 3385 ± 2131 | 0.658 |
| Time spent sitting (min/day) | 446 ± 149 | 386 ± 146 | 0.229 |
| Day of menstrual cycle (# of days) | 16 ± 13 (*n* = 20) | 19 ± 11 (*n* = 12) | 0.391 |
| Mother's level of formal education (au, range 1-7) | 5.7 ± 1.4 | 5.1 ± 1.6 | 0.296 |
| Father's level of formal education (au, range 1-7) | 5.2 ± 1.6 | 5.1 ± 1.5 (*n* = 15) | 0.960 |

**Note:** BMI, body mass index; MET, metabolic equivalent of task.

### Venous Blood Sample

A summary of the venous blood test results is provided in Table 2. There were no differences between groups in Total-C, HDL-C, LDL-C, progesterone, or CRP (*p* > 0.05 for all). However, Black women had lower blood glucose (*p* = 0.013) but higher HbA1c (*p* < 0.001) relative to White women, although the average values for both groups fell within normal healthy ranges. Triglycerides and VLDL-C were higher in the White women (*p* = 0.005 and *p* = 0.004, respectively), but the average values for both groups fell within normal healthy ranges. At the time of testing, serum estradiol was higher in Black women (*p* = 0.043), despite no difference between groups in the number of days into the menstrual cycle (*p* = 0.391; Table 1).

**Table 2.** Venous Blood Sample Results.

| | Black Women<br>mean ± SD (*n*) | White Women<br>mean ± SD (*n*) | *p* value |
| --- | --- | --- | --- |
| Glucose (mg/dl) | 82 ± 6 (19) | 88 ± 5 (14) | 0.013 |
| HbA1c (%) | 5.4 ± 0.3 (19) | 5.1 ± 0.2 (13) | <0.001 |
| Total Cholesterol (mg/dl) | 161 ± 26 (19) | 173 ± 34 (14) | 0.286 |
| Triglycerides (mg/dl) | 51 ± 19 (19) | 86 ± 37 (14) | 0.005 |
| HDL-C (mg/dl) | 64 ± 16 (19) | 58 ± 8 (14) | 0.165 |
| LDL-C (mg/dl) | 86 ± 22 (19) | 99 ± 29 (14) | 0.202 |
| VLDL-C (mg/dl) | 10 ± 3 (19) | 16 ± 6 (14) | 0.004 |
| Estradiol (pg/ml) | 90 ± 82 (16) | 39 ± 40 (11) | 0.043 |
| Progesterone (ng/ml) | 2.7 ± 3.5 (16) | 2.5 ± 4.9 (11) | 0.905 |
| C-Reactive Protein (mg/l) | 1.4 ± 1.7 (19) | 2.9 ± 2.7 (14) | 0.084 |

**Note:** HbA1c, glycated hemoglobin; HDL-C, high-density lipoprotein cholesterol; LDL-C, low-density lipoprotein cholesterol; VLDL-C, very low-density lipoprotein cholesterol.

### Psychosocial Stress

A summary of the psychosocial stress measures is provided in Table 3. There were no differences between groups in adverse childhood experiences or John Henryism endorsement (*p* > 0.05 for both). However, Black women reported greater past week racial/ethnic discrimination (*p* = 0.022). Overall SWS endorsement as well as SWS subscale scores for Black women only are provided in Table 3.

**Table 3.** Psychosocial Stress Exposure & Internalization/Coping.

| | Black Women<br>mean ± SD (*n* = 21) | White Women<br>mean ± SD (*n* = 16) | *p* value |
| --- | --- | --- | --- |
| Adverse Childhood Experiences (#, range 0-10) | 3 ± 2 | 3 ± 3 | 0.389 |
| Past week perceived discrimination (au, range 0-30) | 3.7 ± 4.7 | 1.3 ± 3.5 | 0.022 |
| JHAC12 Score (au, range 12-60) | 46 ± 6 | 48 ± 4 | 0.289 |
| SWS-Overall (au, range 0-105) | 47 ± 15 | - | - |
| SWS-Strength (au, range 0-18) | 11 ± 4 | - | - |
| SWS-Succeed (au, range 0-18) | 12 ± 4 | - | - |
| SWS-Emotions (au, range 0-21) | 12 ± 5 | - | - |
| SWS-Vulnerable (au, range 0-21) | 12 ± 5 | - | - |
| SWS-Help (au, range 0-27) | 12 ± 6 | - | - |

**Note:** JHAC12, John Henryism Active Coping Scale; SWS, Superwoman Schema; Strength, Obligation to present an image of strength; Succeed, Intense motivation to succeed; Emotions, Obligation to suppress emotions; Vulnerable, Resistance to being vulnerable; Help, Obligation to help others.

### Forearm Reactive Hyperemia & Brachial Artery Flow-Mediated Dilation

There were no differences between groups in resting brachial artery blood velocity nor peak blood velocity, absolute change in blood velocity, or percentage change in blood velocity following forearm cuff release (*p* > 0.05 for all; Fig. 1A-D, respectively).

**Figure 1.**
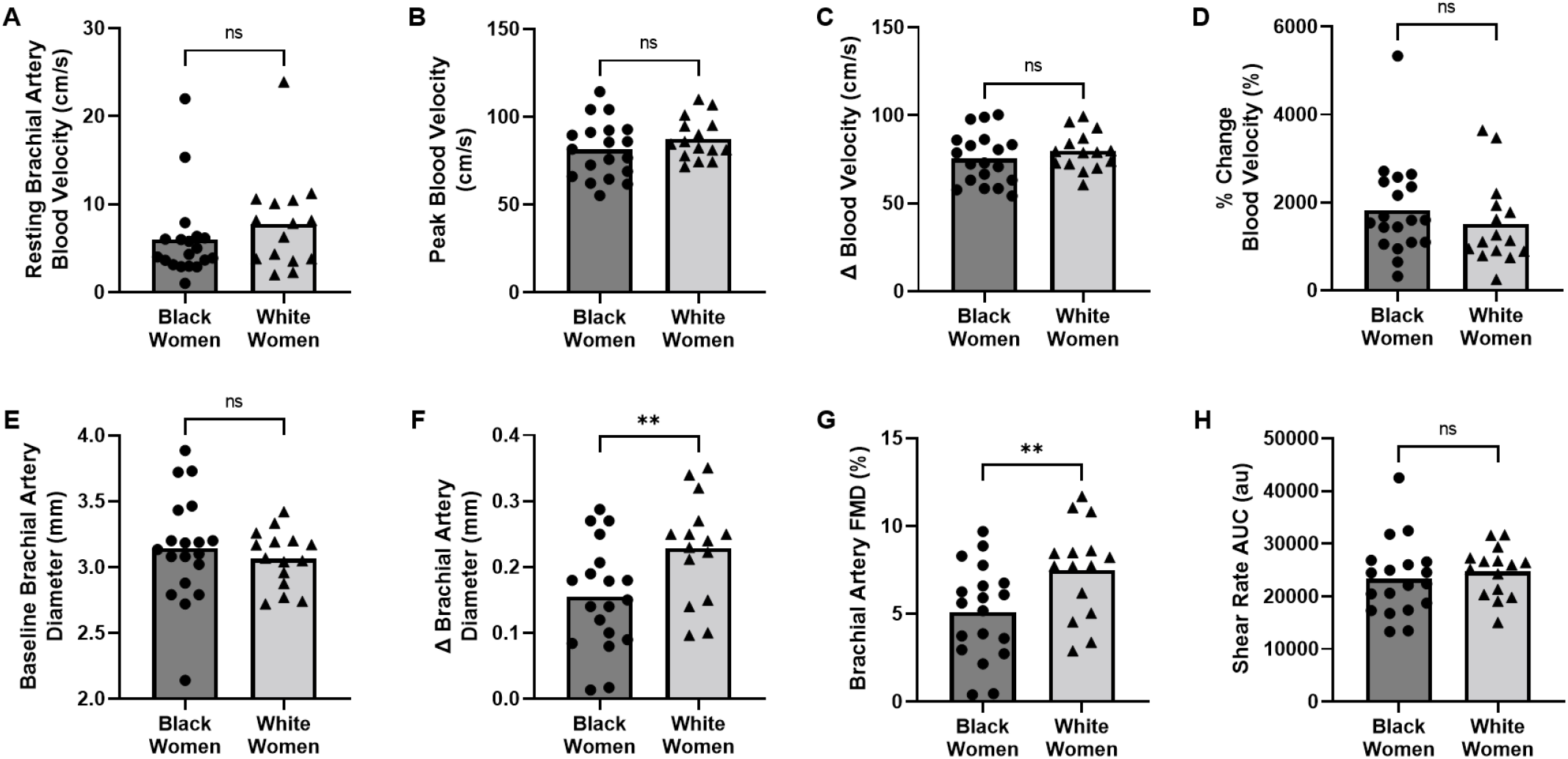
Post-occlusive forearm reactive hyperemia and brachial artery flow-mediated dilation in Black (*n* = 19) and White (*n* = 15) women. A) Resting brachial artery blood velocity. B) Peak brachial artery blood velocity. C) Absolute change in brachial artery blood velocity. D) Percentage change in brachial artery blood velocity. E) Baseline brachial artery diameter. F) Absolute change in brachial artery diameter. G) Percentage change in brachial artery diameter (i.e., flow-mediated dilation). H) Brachial artery shear rate area under the curve (to peak diameter, above baseline). FMD: Flow-mediated dilation. ns: not significant; \**p* < 0.05 between groups. Data are presented individually and as group means.

Baseline brachial artery diameter and shear rate AUC to peak diameter following cuff release were not different between groups (*p* > 0.05 for both; Fig. 1E and Fig. 1H, respectively). Whether expressed as absolute change (Fig. 1F) or percentage change (Fig. 1G) in blood vessel diameter, brachial artery flow-mediated dilation was blunted in Black relative to White women (*p* = 0.006 and *p* = 0.007, respectively), and these results were unaffected when entering shear rate AUC to peak diameter as a covariate in ANCOVA.

### Hypercapnic Cerebrovascular Reactivity

Whether expressed as ΔMCAv/ΔPETCO_2_, ΔCVCi/ΔPETCO_2_, or %CVCi/ΔPETCO_2_, no significant differences between groups were detected in cerebral vascular reactivity to steady-state hypercapnia (*p* > 0.05 for all; Fig. 2 A-C, respectively).

**Figure 2.**
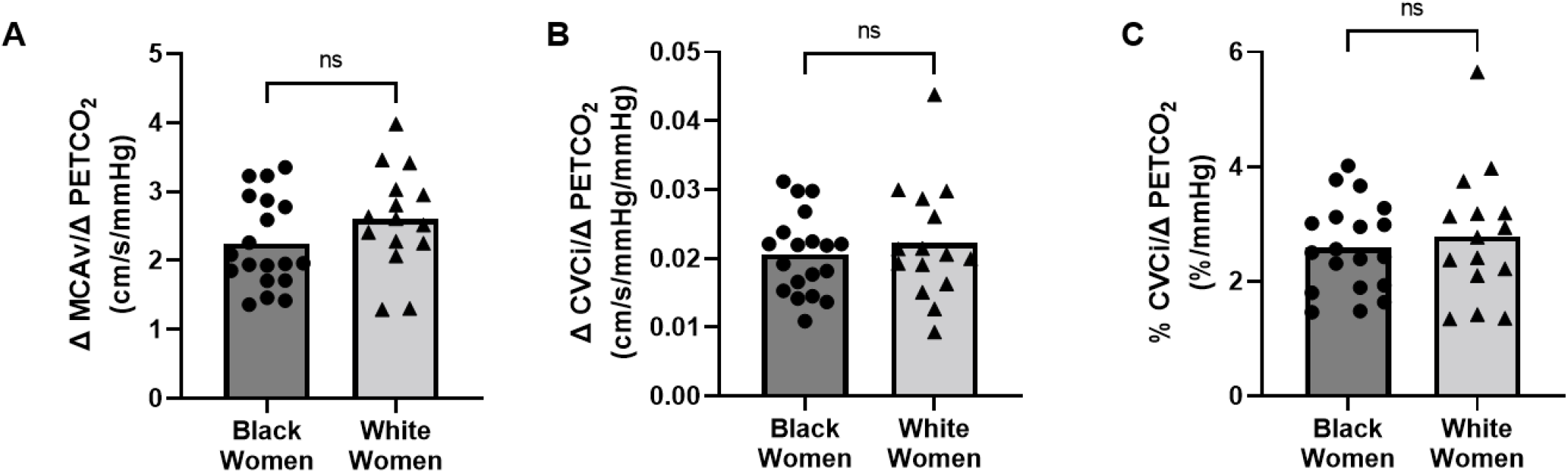
Cerebral vascular reactivity to 6% steady-state hypercapnia in Black (*n* = 19) and White (*n* = 15) women. A) ΔMCAv/ΔPETCO_2_. B) ΔCVCi/ΔPETCO_2_. C) %CVCi/ΔPETCO_2_. MCAv, Middle cerebral artery blood velocity; PETCO_2_, Partial pressure of end-tidal carbon dioxide; CVCi, Cerebrovascular conductance index. ns, not significant. Data are presented individually and as group means.

### Psychosocial Stress Exposure or Internalization/Coping and FMD

There was no relationship between past week discrimination nor adverse childhood experiences and brachial artery FMD in Black (*p* > 0.05 for both; Fig. 3A and 3C, respectively) or White (*p* > 0.05 for both; Fig. 3B and 3D, respectively) women. There was no discernable relationship between Overall SWS (Fig. 4A), SWS-Obligation to Present an Image of Strength (Fig. 4B), SWS-Obligation to Suppress Emotions (Fig. 4D), or SWS-Obligation to Help Others (Fig. 4F) and brachial artery FMD (*p* > 0.05 for all). However, SWS-Intense Motivation to Succeed (Fig. 4C) was and SWS-Resistance to Being Vulnerable (Fig. 4E) tended to be negatively correlated with brachial artery FMD (*r*_s_ = -0.40, *p* = 0.044 and *r*_s_ = -0.37, *p* = 0.057, respectively). In Black women, there was a moderate correlation between John Henryism endorsement and brachial artery FMD (*r*_s_ = -0.50, *p* = 0.014; Fig. 5A). However, in White women, John Henryism endorsement was positively associated with brachial artery FMD (*r*_s_ = 0.53, *p* = 0.042; Fig. 5B).

**Figure 3.**
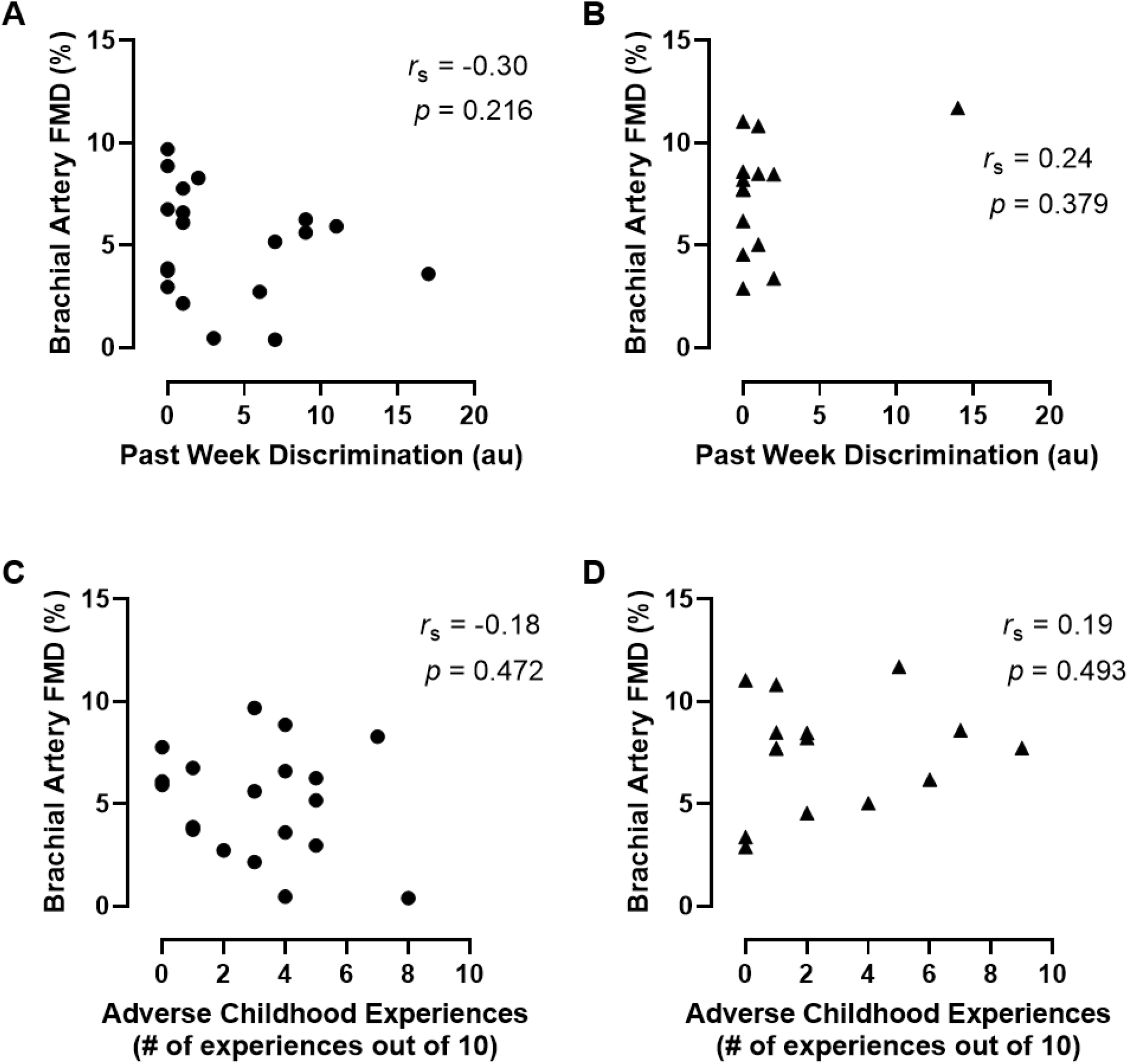
The relationship between Past Week Discrimination (A & B) & Adverse Childhood Experiences (C & D) and brachial artery FMD in Black (⚫, *n* = 19) and White (▲, *n* = 15) women. FMD, flow-mediated dilation.

**Figure 4.**
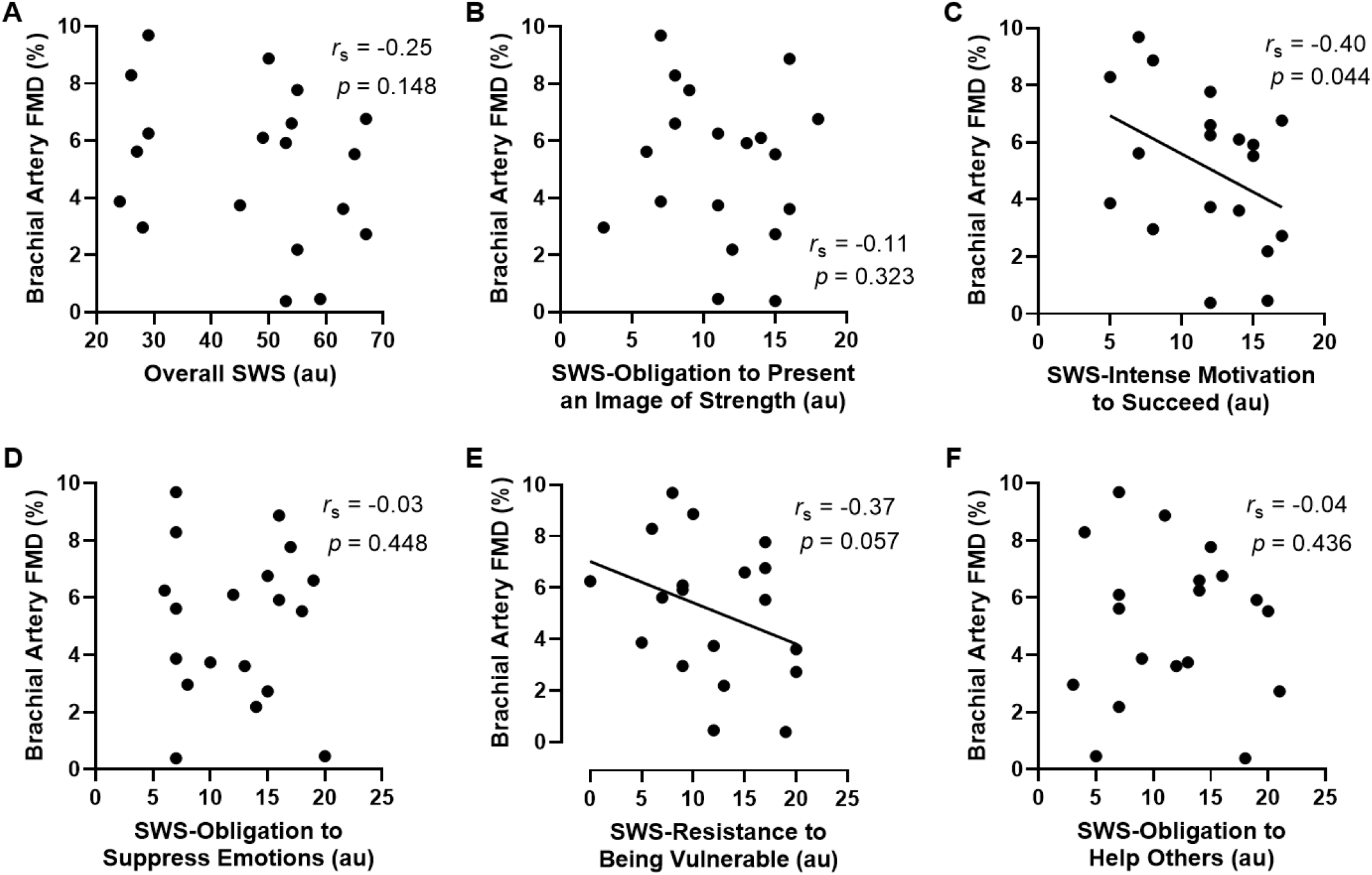
The relationship between Overall SWS (A); SWS-Obligation to Present an Image of Strength (B); SWS-Intense Motivation to Succeed (C); SWS-Obligation to Suppress Emotions (D); SWS-Resistance to Being Vulnerable (E); & SWS-Obligation to Help Others (F) and brachial artery FMD in Black women (*n* = 19). SWS, Superwoman Schema; FMD, flow-mediated dilation.

**Figure 5.**
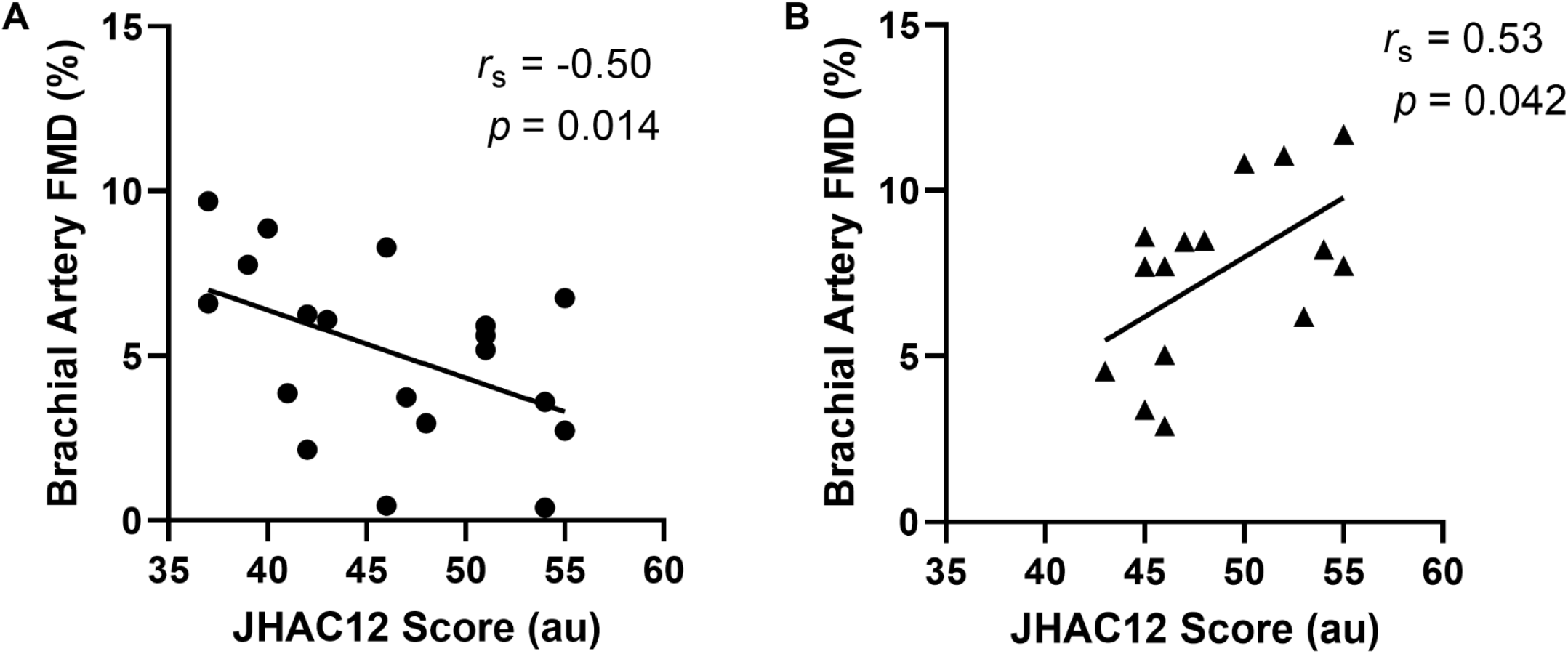
The relationship between endorsement of John Henryism and brachial artery FMD in Black (⚫, *n* = 19) and White (▲, *n* = 15) women. JHAC12, John Henryism Active Coping Scale; FMD, flow-mediated dilation.

## Discussion

The primary findings are that 1) brachial artery (macrovascular) endothelial function but not forearm microvascular nor cerebral vascular function is blunted in young, Black relative to White women and 2) among Black women, there are stronger relationships between indices of psychosocial stress internalization/coping techniques and brachial artery FMD than psychosocial stress exposures alone and FMD. To our knowledge, this is the first study to investigate the relationship between various indices of psychosocial stress exposure and internalization/coping techniques and novel indices of peripheral and cerebral vascular function in Black women, a population that faces heightened CVD and stroke risk. Importantly, the National Institutes of Health and the Center for Disease Control and Prevention have substantially increased efforts to address social determinants of health and structural racism (46, 47). Additionally, a recent Presidential Advisory from the American Heart Association called for accelerated research into the joint effects of multiple domains of racism on health (48). Accordingly, by investigating the potential influence of psychosocial stress exposure and internalization/coping techniques on indices of peripheral and cerebral vascular function, the present study provides new knowledge regarding the relationship between interpersonal racism and cardiovascular health.

### Brachial Artery Flow-Mediated Dilation

Brachial artery FMD testing is a common approach for assessing endothelial function (49) and is predictive of CVD morbidity and mortality (2-5). Here, we demonstrated that young, otherwise healthy Black women have blunted brachial artery endothelial function. Few studies have investigated differences by race/ethnicity in conduit artery endothelial function in this manner. However, our data are consistent with and extend the findings of those from the available studies by Campia et al. (12), Perregaux et al. (50), and D’Agata et al. (7), which all demonstrate blunted FMD in Black relative to White women. Although our approach was generally similar to those studies, there are some differences to consider. Campia et al. (12) studied an older (∼40 yr) cohort whereas our participants were ∼20-25 yr; therefore, the present findings provide new data in a younger population. Additionally, the current data are somewhat conflicting with the conclusions of the study from D’Agata et al. (7) who reported blunted brachial artery FMD in a cohort of Black women (7). However, in their study, differences were abolished after normalizing the raw FMD values to shear rate AUC (7), leading them to conclude that peripheral macrovascular function was preserved in Black relative to White women. We, however, accounted for shear rate AUC by reporting/examining it (and found no differences) as is recommended by the current guidelines for FMD testing (42). Accordingly, our data provide new evidence that brachial artery FMD is blunted in young, otherwise health Black women. Potential mechanisms for this disparity in vascular endothelial function are discussed below.

### Forearm Reactive Hyperemia

Post-occlusive RH testing is commonly utilized to assess microvascular function (51), and blunted responses are predictive of future CVD risk (2). In the present study, there were no differences in forearm reactive hyperemia between Black and White women. This finding is contrary to the conclusions of the study by D’Agata et al. (7), where they indicated that forearm microvascular function was blunted in Black women. However, this conclusion was based on RH indexed as total flow AUC, which is confounded by changes in brachial artery diameter over the course of the recovery period. Consistent with our data, D’Agata et al. (7) reported no differences between Black and White women in peak hyperemia, which appears to be a better indicator of truly “reactive” hyperemia as opposed to “responsive” hyperemia, as proposed by Rosenberry & Nelson (51). Some (52, 53) but not all (12, 50) studies investigating differences in reactive hyperemia by race/ethnicity have demonstrated that Black individuals have blunted microvascular function (52, 53). Although performed in a middle-aged cohort, the study by Morris et al. (53) was designed to investigate race x sex differences and indicated that there were no differences between Black and White women, while the study by Ranadive et al. (52) was not designed to investigate race x sex differences. Therefore, our finding of preserved reactive hyperemia, as indexed by peak hyperemia, in Black relative to White women is consistent with that of D’Agata et al. (7) and Morris et al. (53); however larger investigations are needed to completely understand whether there are differences in forearm microvascular function by race/ethnicity, particularly in young, otherwise health Black and White women

### Hypercapnic Cerebrovascular Reactivity

Cerebral vascular reactivity serves as an index of cerebral vascular function/health and is predictive of all-cause, cardiovascular, and non-cardiovascular mortality (6, 54). To our knowledge, this is the first study to specifically investigate cerebral vascular reactivity to hypercapnia in young Black and White women. We demonstrated that there were no differences between groups regardless of the metric used to express CVR. Previous data from our lab indicate that young, Black men and women have blunted hypercapnic CVR (10, 14); however, these studies were not designed to investigate race x sex differences. Accordingly, the previously observed race/ethnicity-related differences in CVR may be primarily driven by blunted CVR in Black men. In support of this, Akins et al. (55) recently demonstrated that CVR was significantly blunted in Black men relative to Black women. There are numerous possibilities to explain these potential race x sex differences in CVR. Given that all the women participants in the studies (10, 14, 55) from our lab are premenopausal, the cerebral vasculature in Black women may be protected in part by estrogen, which appears to preserve cerebral vascular function (56, 57). More longitudinal work that leverages the SDOH framework (16) is needed to better understand how Black women transition from preserved CVR in young adulthood to having a twofold higher stroke prevalence relative to White women (1).

### Psychosocial Stress Exposure or Internalization/Coping and FMD

Given that Black women exhibited blunted conduit artery endothelial function in the present study, we investigated whether there were differential relationships between brachial artery FMD and psychosocial stress exposure vs. internalization/coping. We found that there was no discernible relationship between psychosocial stress exposure, as indexed by past week discrimination and adverse childhood experiences, and FMD among Black or White women. However, among Black women, the relationship between FMD and endorsement of Overall SWS as well as the SWS subscales varied from no relation to weak-to-moderate. The subscales that either were correlated with or tended to correlate with FMD were SWS-Intense Motivation to Succeed and SWS-Resistance to Being Vulnerable respectively. In comparison, John Henryism endorsement exhibited a moderate inverse relationship with FMD among Black women while, interestingly, a moderate positive relationship between John Henryism endorsement and FMD was observed in White women. Taken together, these data indicate that psychosocial stress exposure alone is not related to brachial artery endothelial function in young, Black or White women. However, depending on the metric used, psychosocial stress internalization/coping techniques may exhibit a moderate inverse relationship with brachial artery endothelial function in Black women. Although the finding of a positive relationship between John Henryism and FMD in White women was not anticipated, this is not unprecedented (58), and it is possible that John Henryism is associated with health-promoting behaviors in some groups. For example, Bild et al. (59) demonstrated that John Henryism was positively associated with higher physical activity among White women (59), which itself has numerous positive effects on cardiovascular health/function (60). Nevertheless, these data set the stage for more comprehensive investigations aiming to better understand relationships between various domains of psychosocial stress and cardiovascular health and function, particularly in Black women. In general, the literature suggests that psychosocial stress exposure is predictive of cardiovascular health (61); however, the additive power of internalization and coping techniques, within a broader context of structural racism, is immense (26-29). Therefore, in our relatively young cohort of Black women, it appears that one’s coping techniques may have a greater contribution to alterations in endothelial function than exposure to psychosocial stressors alone. Research regarding the mechanisms that link psychosocial stress exposure and internalization/coping to cardiovascular function and outcomes is an area of heavy interest (29, 58, 62-64). Although numerous theories and hypotheses have been proposed and continue to be tested (21), the prevailing hypothesis is that psychosocial stress gets “under the skin” to promote numerous physiological changes (20, 65). In the short term, such changes (i.e., allostasis), including increased sympathetic nervous system activity and inflammation, can be protective. However, when upregulated over a sustained period, these changes are pathophysiological (i.e., allostatic overload) (20, 21). Our data generally support this hypothesis but only among Black women; however, longitudinal investigations will be required to parse out the influence of time on the magnitude of the exposure- vs. internalization/coping-endothelial function relationship in this and other populations who face greater psychosocial stress.

### Strengths and Limitations

The participants in this study had similar body composition, physical activity and sedentary levels, and resting heart rate and blood pressure (Table 1). This strengthens the interpretability of our findings by reducing the influence of confounding variables. This study was also strengthened by the assessment of a multitude of well-validated vascular health combined with various psychosocial outcomes. Few studies have performed such comprehensive vascular health assessments alongside psychosocial measures in at-risk populations.

Our approach of favoring complex tests of vascular function necessitated a relatively small sample size, which is a limitation of the current investigation. Accordingly, these findings set the stage for larger cross-sectional and longitudinal studies. Furthermore, our sample consisted mainly of college students, particularly in the Black women group. This reduces the overall socioeconomic diversity and thus the diversity of exposure to psychosocial stressors within the cohort and therefore the findings of the present investigation may not be representative of young, Black women across the Dallas-Fort Worth metroplex let alone the United States. Despite this narrower range of socioeconomic diversity, we still observed some inverse relationships between indices of vascular health and psychosocial stress internalization/coping. Therefore, based on our current findings, it could be hypothesized that the negative effect of these psychosocial stressors would only be further amplified with a more socioeconomically diverse cohort. Future work should build on the current investigation by conducting multi-site trials that include aggressive yet sensitive recruitment strategies capable of incorporating a more representative sample as the current study may overrepresent Black women of higher seriocomic status, which is typically associated with attending college. Regarding the overall younger age of our cohorts, we intentionally studied a young, healthy population because our focus is on the prevention of CVD by identifying early, subclinical markers of future CVD risk.

### Conclusion

To our knowledge, this is the first study to investigate peripheral and cerebral vascular function alongside novel indices of psychosocial stress exposure and internalization/coping techniques in young, Black women. Our data indicate that peripheral microvascular and cerebral vascular function are preserved in young Black women, but conduit artery endothelial function is blunted. Furthermore, we demonstrated that blunted conduit artery endothelial function exhibited a stronger relationship with psychosocial stress internalization/coping techniques relative to exposure to psychosocial stressors alone in Black women. Collectively, these findings indicate that elevated CVD risk in Black women may be partially attributable to internalization of psychosocial stressors as well as maladaptive coping techniques.

## Data Availability

These data supporting the findings and conclusions of this study are available, upon reasonable request, from the corresponding author on request

## Author statements

### Availability of data

The raw data are available upon request.

## Acknowledgments

The authors would like to thank the participants for their time and effort in this study.

